# Performance of blood biomarkers in internal jugular vein for Alzheimer disease pathologies: the Delta Study

**DOI:** 10.1101/2025.03.09.25323608

**Authors:** Jun Wang, Dong-Yu Fan, Shan Huang, Chang Liu, Pei-Wen Zhao, Nan Wu, Xiao-Lin Gao, Qing-Zhi Wang, Yanli Li, Bai Liu, Yuan-Ye Ma, Rong-Chang Zhao, Yu-Peng Zhu, Qiong-Yan Li, Xiao-Yu Liu, Xiao Chen, Yu-Jie Lai, Fan Zeng, Yu-Hui Liu, Xian-Le Bu, Tengfei Guo, Yi-Tang, Jin-Tai Yu, Colin L. Masters, Junhong Guo, Qingxiang Mao, Jing Yang, Yan-Jiang Wang, the Translational Biomarker Research of AgeIng and Neurodegeneration (TBRAIN)

**Author notes:** Correspondence to: Yan-Jiang Wang, 10 Changjiang Branch Road, Daping, Yuzhong district, Chongqing 400042, China., Jing Yang, No. 1 Eastern Jianshe Road, The First Affiliated Hospital of Zhengzhou University, Zhengzhou University, Zhengzhou 450052, China., Qingxiang Mao, 10 Changjiang Branch Road, Daping, Yuzhong district, Chongqing 400042, China., Junhong Guo, No.85 Jiefang South Street, First Hospital of Shanxi Medical University, Taiyuan 030012, China. These authors contributed equally to this work (Jun Wang, Dong-Yu Fan, Shan Huang).

## Abstract

**Background:** Systemic factors confound blood tests for the diagnosis of Alzheimer disease (AD). The Delta study explored whether blood biomarkers from the vein proximal to the brain perform better in detecting cerebral AD pathologies.

**Methods:** Blood was collected from the internal jugular vein (IJV) and median cubital vein (MCV) in the discovery (n=371) and validation (n=92) cohorts. AD biomarkers were measured with Lumipulse G and Simoa methods. Aβ and tau PET imaging and cerebrospinal fluid (CSF) biomarkers were used to evaluate brain pathologies.

**Results:** The levels of Aβ42, Aβ40, p-tau217, p-tau181, GFAP and NfL were higher in the IJV than MCV and highly correlated between the two sites. IJV-Aβ42/40 had stronger correlations with Aβ PET Centiloids and tau PET meta-temporal SUVR than MCV-Aβ42/40. In detecting cerebral Aβ positivity, IJV-Aβ42/40 demonstrated a significantly higher accuracy (79.9% to 92.9% vs. 72.4% to 88.8%) and a lower percentage of uncertain individuals (17.8% to 54.5% vs. 31.3% to 70.1%) than MCV-Aβ42/40. Moreover, the diagnostic accuracy of Lumipulse G IJV-Aβ42/40 (88.2% to 92.9%) was statistically equivalent to that of MCV-p-tau217 (90.2% to 94.3%), although the intermediate percentage of IJV-Aβ42/40 was higher (17.8% to 34.0% vs. 0.7% to 17.5%) than MCV-p-tau217. These findings were verified in the validation cohort.

**Discussion:** IJV-Aβ42/40 performs better than MCV-Aβ42/40 in detecting cerebral AD pathologies, offering a novel perspective to reduce the impacts of systemic factors and comorbidities on blood tests.

## Introduction

Senile plaques composed of amyloid-beta (Aβ) peptides in the brain are a specific pathology of Alzheimer’s disease (AD). Blood biomarkers reflecting cerebral Aβ pathology, such as Aβ and p-tau, have recently been incorporated into updated diagnostic criteria for AD^1,2^. While blood phosphorylated-tau (p-tau) proteins, including p-tau181 and p-tau217, demonstrate high performance in identifying brain Aβ pathology^3,4^, they are neither direct nor specific biomarkers for cerebral Aβ deposition. This limitation arises because p-tau levels are also elevated in other neurodegenerative diseases, such as neuronal intranuclear inclusion disease (NIID) and Creutzfeldt–Jakob disease (CJD)^5,6^. Furthermore, increases in blood p-tau217 in AD patients are apparent until Braak stage IV or later, diminishing its utility for early diagnosis^7^. The blood Aβ42/40 ratio, though theoretically specific to cerebral Aβ pathology, has shown suboptimal diagnostic performance. This is attributed to its modest inter-group fold-change (reduced by only 8%–15% in AD patients) and substantial variability influenced by systemic factors or comorbidities such as chronic kidney disease (CKD), myocardial infarction, and stroke ^7–10^.

Biomarkers derived from venous blood proximal to the brain may increase diagnostic accuracy, as biomarkers at this site are less diluted by systemic circulation and are minimally confounded by peripheral metabolic processes. To explore this hypothesis, we conducted a multicentre clinical study comparing the diagnostic performance of AD biomarkers in blood collected from the internal jugular vein (IJV), the anatomical site closest to the brain, and the median cubital vein (MCV), the conventional site for blood sampling.

## Methods

### Study participants

The study design is shown in **Fig. 1**. This multicentre study included two independent cohorts (n=463). The discovery cohort (n=371) of the Chongqing Ageing & Dementia Study (CADS)^12^ was from Daping Hospital (Chongqing, China) from January 2019 to November 2024. The validation cohort were from the First Affiliated Hospital of Zhengzhou University (Henan, China) and the First Hospital of Shanxi Medical University (Shanxi, China) from January 2024 to December 2024, and from Daping Hospital from December 2024 to January 2025. Participants who had complaints of cognitive decline or consulting for the risk of developing AD and were willing to participate in this study were consecutively enrolled. Patients with severe diseases who could not comply with complete PET scans or blood sampling or who refused to participate in this study were excluded (eMethods). The diagnosis of AD was according to the 2024 Alzheimer’s Association criteria and the 2024 Clinical-Biological Construct^1,2^.

**Figure 1.**
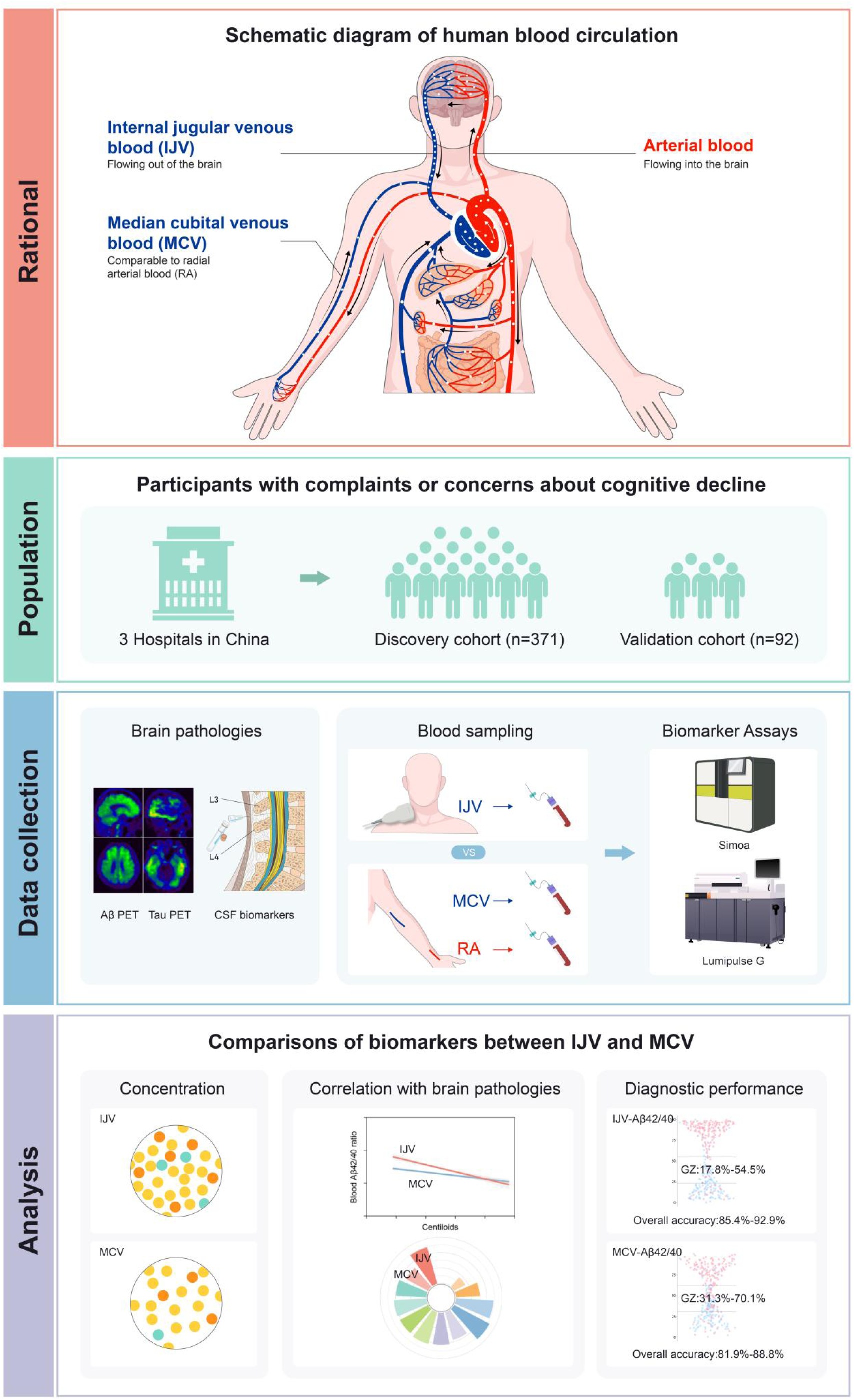
Study design. In human blood circulation, the median cubital vein (MCV) is the routine site for venous blood collection. The MCV blood is transformed by the exchange of the arterial blood through the capillaries of the unilateral upper limb. Meanwhile, arterial blood flows into the brain, passing through the exchange of capillaries and carrying brain-derived molecules out of the brain. The internal jugular vein (IJV), which contains brain-derived proteins not yet metabolized by the periphery, is closest to the brain and is clinically safe and easy to draw blood from.

This study was approved by the Institutional Review Board of the three hospitals, and all participants and their caregivers provided informed consent (ClinicalTrials.gov number, NCT06754254.).

### Blood and CSF sampling and processing

Daping Hospital is a member of the Alzheimer’s Association Quality Control Program for CSF and blood biomarkers and led this study. As members of the Translational Biomarker Research of AgeIng and Neurodegeneration Consortium (TBRAIN), all three research centres adopted the same standard operating procedure (SOP) of biofluid collection and processing^13^. Fasting blood from the IJV and MCV was collected using the same blood collection device within 1 hour to ensure the comparability of biomarker levels among different circulation sites and to avoid the influence of operating procedures. In the radial artery (RA) subcohort of the CADS, RA, MCV and IJV blood were collected within 10 minutes. IJV blood sampling was completed by experienced anaesthesiologists under ultrasound guidance, and was well-tolerated by all the participants. Plasma was isolated and stored at −80 °C until use (eMethods).

### Measurements of blood and CSF biomarkers

Plasma biomarkers were measured on two fully automated platforms in the laboratory at one centre (Daping Hospital): (1) plasma Aβ42, Aβ40, p-tau181, and p-tau217 using the Lumipulse G1200 platform and (2) plasma Aβ42, Aβ40, p-tau217, GFAP, and NfL using a single-molecule array [Simoa] on the HD-X analyser. MCV, IJV and RA blood samples were measured simultaneously.

CSF biomarkers (Aβ42, Aβ40 and p-tau181) were measured on the Lumipulse G1200 platform, which has achieved FDA approval for the detection of Aβ plaques associated with AD.

### PET acquisition and analysis

Aβ PET is performed with the [^11^C]Pittsburgh Compound B (PiB) or [^18^F]florbetapir (AV45) tracer, and tau PET is performed with the [^18^F]MK-6240 tracer. Aβ and tau PET images were visually analysed using a dichotomous negative/positive PET read by two experienced nuclear medicine physicians^14^. Any disagreement between the two readers was resolved by consulting a third senior assessor. Meanwhile, Aβ and tau PET images in the CADS cohort were quantified using cortical standard centiloids (CLs) and meta-temporal standard uptake value ratio (SUVR) using the CapAIBL platform^15^, respectively.

### Statistical analysis

Between-group differences were analysed using the paired t-test, Wilcoxon matched-pairs signed rank test, Mann‒Whitney U test or Kruskal‒Wallis test, as appropriate. Correlation coefficients were compared using the Fisher Z-transformation. The slopes of the two regression lines were compared using the F-test. Univariate linear regression models with the coefficient of determination (R^2^) were used to quantify the contributions of brain pathologies to blood biomarkers.

The single- and two-cutoff approaches were used to evaluate the performance of IJV and MCV biomarkers in predicting brain pathologies. The single-cutoff approach was based on the maximum Youden index using the receiver operating characteristic (ROC) curve. In the two-cutoff approach, the lower threshold was obtained by maximizing the specificity with the sensitivity fixed at 90% or 95%, respectively, whereas the upper threshold was by maximizing the sensitivity with the specificity fixed at 90% or 95%. Diagnostic metrics (including area under the curve [AUC], accuracy, sensitivity, specificity, positive prediction value [PPV], and negative prediction value [NPV]) were calculated as the mean with 95% CI of the bootstrapped resample (n = 1,000). The differences between the two biomarkers were considered equivalent when the 95% CI range included zero.

All analyses were performed using R studio version 4.2.2 (R Project for Statistical Computing), with a 2-sided α of 0.05. Figures were generated using the graphpad prism version 10.0 and R studio.

## Results

### Participant characteristics

The discovery cohort comprised 371 participants with a mean age (standard deviation [SD]) of 65.4 (9.6) years, including 203 females (54.7%) and 130 *APOE ε4* carriers (35.0%). Among these participants, 323 underwent Aβ PET examination with 149 (46.1%) showing positive results, while 269 completed tau PET scans and 234 underwent CSF tests. The RA subcohort included 65 participants, with a mean age of 64.6 (8.9) years. The validation cohort included 92 participants with a mean age of 62.0 (9.1) years. Among them, 49 of 89 (55.1%) had positive Aβ status. The characteristics of the populations are listed in **Table 1**.

**Table 1.**
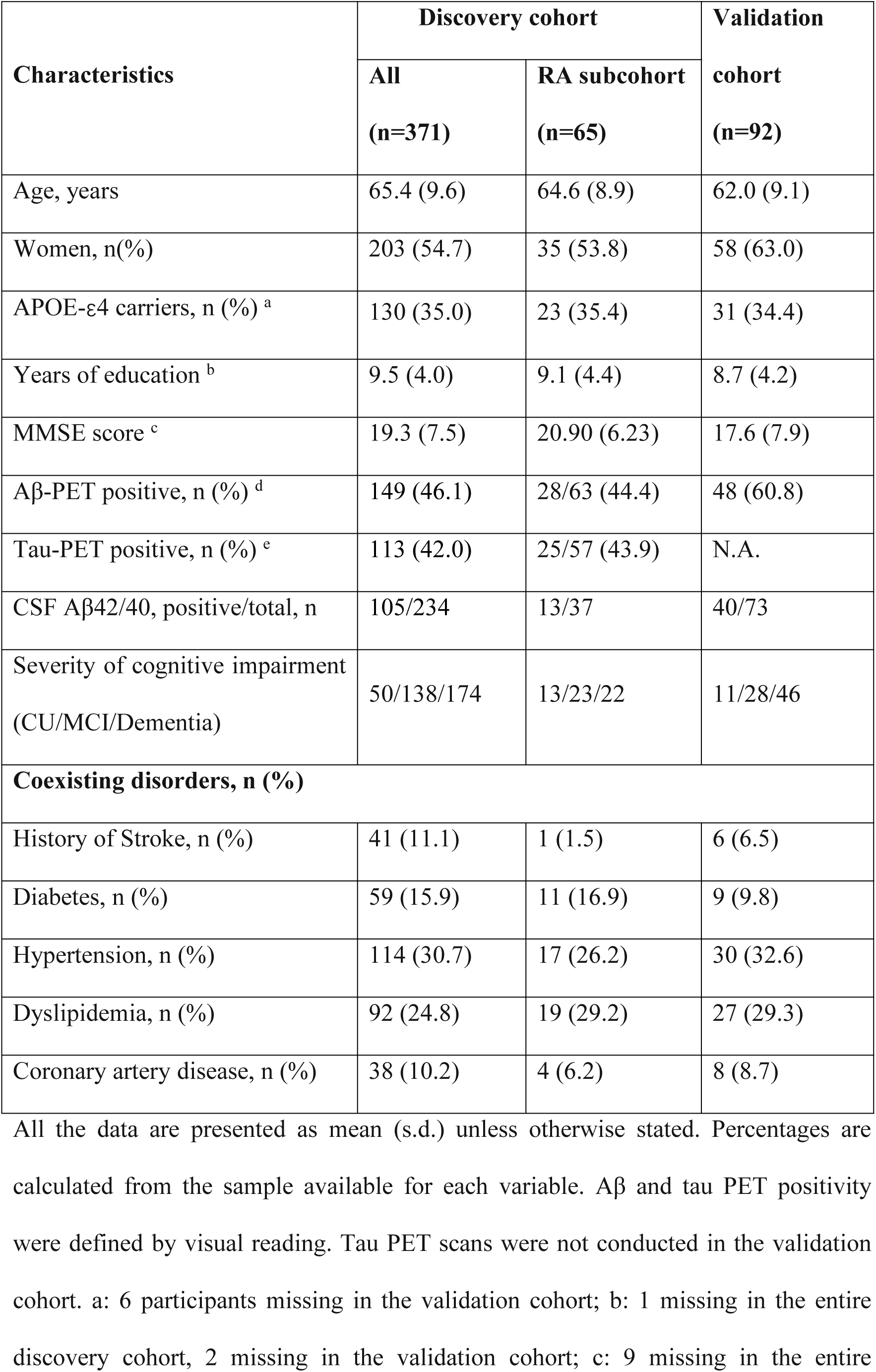
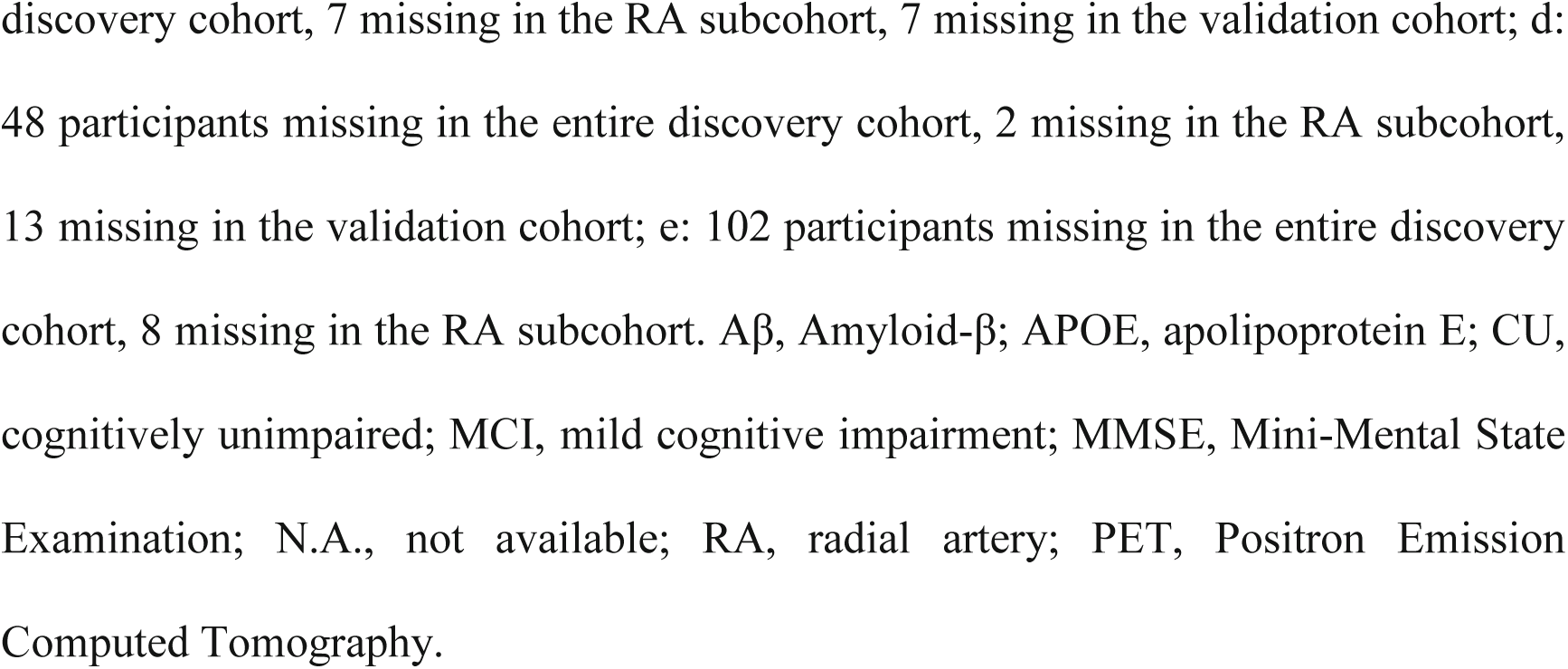
Characteristics of participants.

| Characteristics | Discovery cohort |  | Validation cohort<br>(n=92) |
| --- | --- | --- | --- |
|  | All<br>(n=371) | RA subcohort<br>(n=65) |  |
| Age, years | 65.4 (9.6) | 64.6 (8.9) | 62.0 (9.1) |
| Women, n(%) | 203 (54.7) | 35 (53.8) | 58 (63.0) |
| APOE-ε4 carriers, n (%) <sup>a</sup> | 130 (35.0) | 23 (35.4) | 31 (34.4) |
| Years of education <sup>b</sup> | 9.5 (4.0) | 9.1 (4.4) | 8.7 (4.2) |
| MMSE score <sup>c</sup> | 19.3 (7.5) | 20.90 (6.23) | 17.6 (7.9) |
| Aβ-PET positive, n (%) <sup>d</sup> | 149 (46.1) | 28/63 (44.4) | 48 (60.8) |
| Tau-PET positive, n (%) <sup>e</sup> | 113 (42.0) | 25/57 (43.9) | N.A. |
| CSF Aβ42/40, positive/total, n | 105/234 | 13/37 | 40/73 |
| Severity of cognitive impairment<br>(CU/MCI/Dementia) | 50/138/174 | 13/23/22 | 11/28/46 |
| **Coexisting disorders, n (%)** |  |  |  |
| History of Stroke, n (%) | 41 (11.1) | 1 (1.5) | 6 (6.5) |
| Diabetes, n (%) | 59 (15.9) | 11 (16.9) | 9 (9.8) |
| Hypertension, n (%) | 114 (30.7) | 17 (26.2) | 30 (32.6) |
| Dyslipidemia, n (%) | 92 (24.8) | 19 (29.2) | 27 (29.3) |
| Coronary artery disease, n (%) | 38 (10.2) | 4 (6.2) | 8 (8.7) |

All the data are presented as mean (s.d.) unless otherwise stated. Percentages are calculated from the sample available for each variable. Aβ and tau PET positivity were defined by visual reading. Tau PET scans were not conducted in the validation cohort. a: 6 participants missing in the validation cohort; b: 1 missing in the entire discovery cohort, 2 missing in the validation cohort; c: 9 missing in the entire
discovery cohort, 7 missing in the RA subcohort, 7 missing in the validation cohort; d: 48 participants missing in the entire discovery cohort, 2 missing in the RA subcohort, 13 missing in the validation cohort; e: 102 participants missing in the entire discovery cohort, 8 missing in the RA subcohort. Aβ , Amyloid-β ; APOE, apolipoprotein E; CU, cognitively unimpaired; MCI, mild cognitive impairment; MMSE, Mini-Mental State Examination; N.A., not available; RA, radial artery; PET, Positron Emission Computed Tomography.

### Blood biomarker levels at different circulation sites

The levels of Aβ42, Aβ40 and p-tau217 measured using the two methods demonstrated strong intermethod correlations (**Fig. S1**). All AD biomarker levels were higher in the IJV than in the MCV (p <0.001; **Fig. 2A and C)**, with robust intersite correlations (Spearman ρ=0.74–0.99; **Fig. S2A and B**).

**Figure 2.**
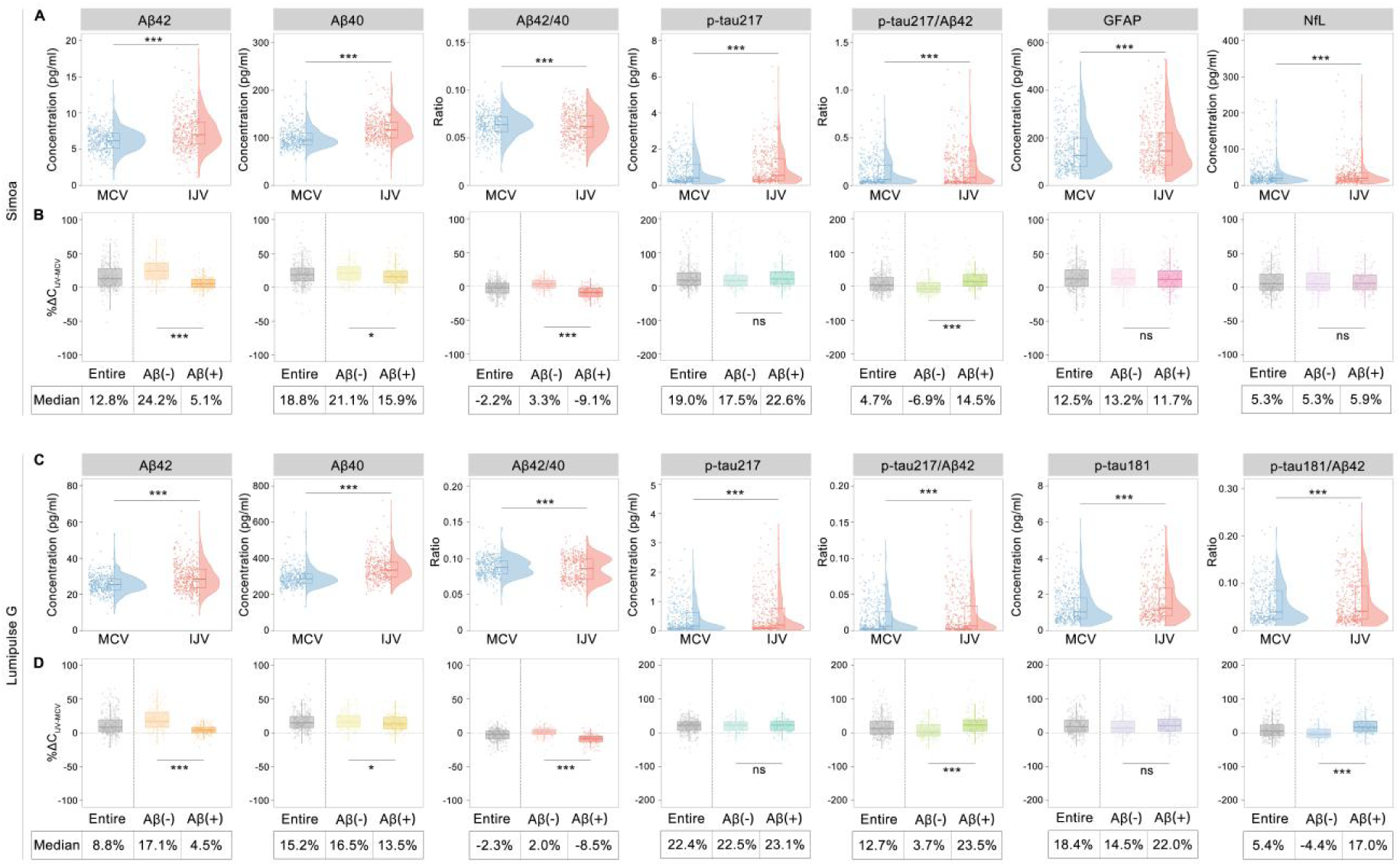
Comparisons of blood biomarker levels between the MCV and IJV. **(A,C)** Levels of AD biomarkers in the IJV and MCV. **(B,D)** Differences in blood biomarker levels between the MCV and IJV in the entire cohort and Aβ-positive and Aβ--negative subgroups. %ΔC_IJV-MCV_ of biomarkers was calculated as: (C_IJV_-C_MCV_)/C_MCV_*100%. Aβ-positive (Aβ+) or Aβ-negative (Aβ-) status was defined by Aβ PET or CSF Aβ42/40 ratio. IJV, internal jugular vein; MCV, median cubital vein. C_IJV_, concentration in the IJV; C_MCV_, concentration in the MCV. Comparisons of biomarker levels between the MCV and IJV were performed using paired t tests for Aβ42, Aβ40 and Aβ42/40, and using Wilcoxon matched-pairs signed rank test for others. *p<0.05; **p<0.01; ***p<0.001; ns, not significant.

The concentration gradient between the IJV and RA represents the net efflux of biomarkers from the brain. In the RA subcohort, minimal yet highly correlated biomarker differences were observed between RA and MCV (**eResults and Fig. S3**). For practical feasibility, MCV biomarker levels were adopted as RA proxies to compute the IJV–MCV differential (ΔC_IJV-MCV_), substituting for the theoretical ΔC_IJV-RA_ in efflux calculations. The intersite concentration differences (%Δ=C_IJV_-_MCV_/C_MCV_×100%) displayed biomarker-specific patterns: p-tau217 presented the highest percentage differential (median %Δp-tau217: 19.0%–22.4%), whereas Aβ42 exhibited the lowest (median %ΔAβ42: 8.8%–12.8%) among the core AD biomarkers. None-core biomarkers demonstrated smaller gradients (%ΔGFAP: 12.5%; %ΔNfL: 5.3%). Biomarker ratios displayed reduced variability compared to individual analytes (Aβ42/40: −2.3%– −2.2%; p-tau217/Aβ42: 4.7%–12.7%; p-tau181/Aβ42: 5.4%; **Fig. 2B and D)**.

Stratified analysis revealed marked reductions in the Aβ42 and Aβ40 gradients in Aβ(+) versus Aβ(−) subgroups (median %ΔAβ42: 4.5–5.1% vs. 17.1–24.2%; %ΔAβ40: 13.5–15.9% vs. 16.5–21.1%; p < 0.05; **Fig. 2B and D**). No significant intergroup differences were observed for p-tau isoforms (%Δp-tau217, %Δp-tau181; p > 0.05) or other biomarkers (%ΔGFAP, %ΔNfL; p > 0.05).

The above findings were verified in the validation cohort (**Figs. S2 and S4**).

### Correlations between biomarkers and cerebral pathologies

In the discovery cohort, IJV-Aβ42/40 exhibited stronger correlations with cerebral Aβ burden (as quantified by cortical average Centiloids on Aβ-PET) than MCV-Aβ42/40 (Spearman ρ: −0.608– −0.487 vs. −0.485– −0.328, p<0.05). While p-tau217 and p-tau181 presented comparable correlation strengths between the IJV and MCV with Centiloids, linear regression analyses revealed steeper slopes for IJV p-tau, suggesting greater Aβ-associated p-tau level changes in the IJV than in the MCV (**Fig. 3A**). No significant intervenous differences were observed for the correlations of GFAP or NfL with Centiloids (**Fig.S5A**). Notably, in the Aβ-PET(+) populations, IJV-Aβ42/40 and ΔAβ42/40, rather than MCV-Aβ42/40, were associated with Centiloids (**Fig. S6**).

**Figure 3.**
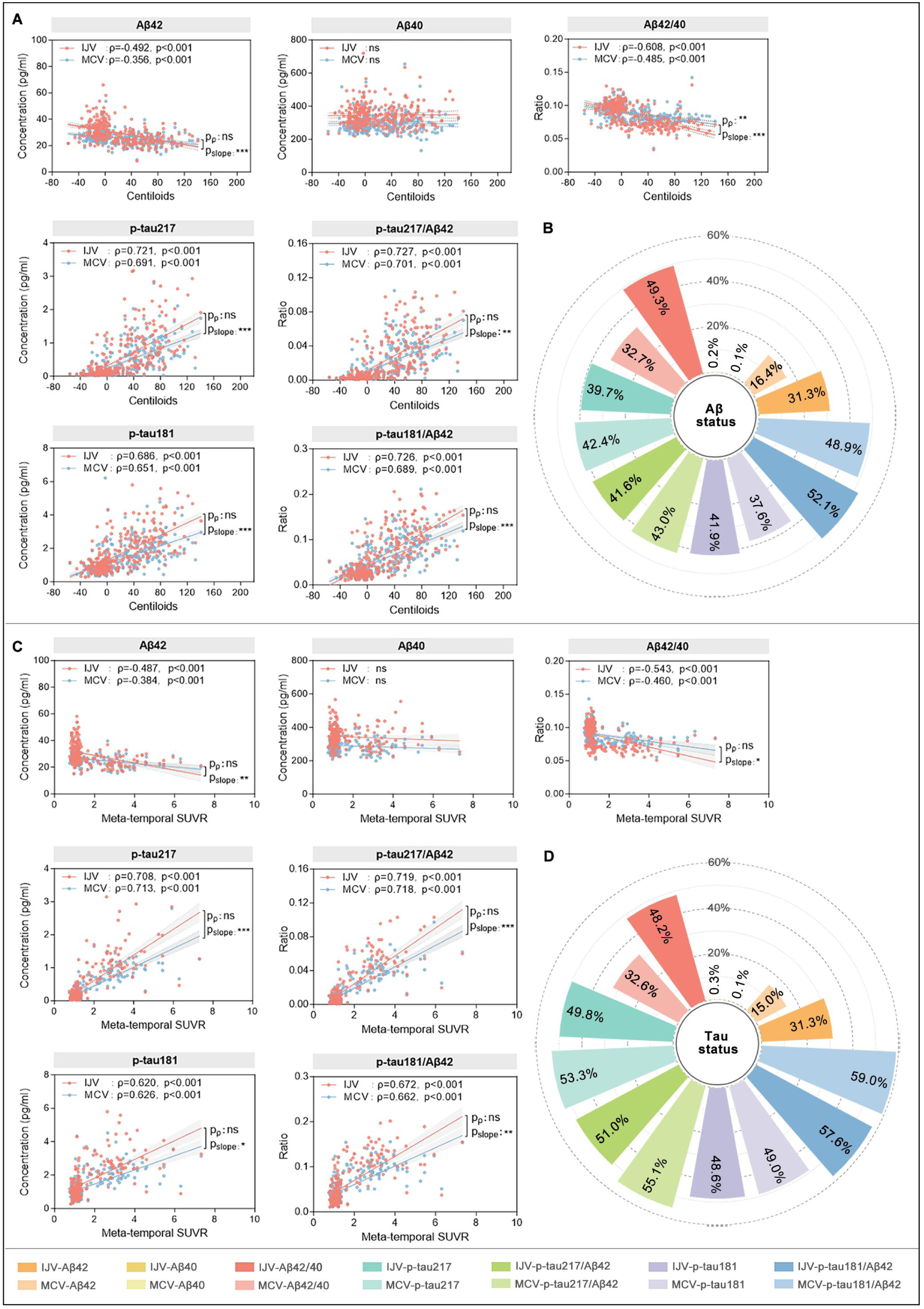
Correlations of the IJV and MCV biomarkers measured on the Lumipulse G platform with brain pathologies. **(A)** Correlations of the IJV and MCV biomarkers with the brain Aβ burden measured by Aβ-PET Centiloids. **(B)** Contribution of the brain Aβ status to the IJV and MCV biomarkers. Aβ status was defined by a visual read of Aβ PET imaging or CSF Aβ42/40 ratio. **(C)** Correlations of IJV and MCV biomarkers with brain tau pathology as measured by tau-PET meta-temporal SUVR. **(D)** Contribution of brain tau status to IJV and MCV biomarkers. Tau status was defined by a visual read of Tau PET imaging. CSF, cerebrospinal fluid; IJV, internal jugular venous blood; MCV, medial cubital venous blood; PET, positron emission tomography; SUVR, standardized uptake value ratio. p_ρ_, p value for Spearman correlation coefficient ρ; p_slope_, p value for slope of the fitted linear. *p<0.05; **p<0.01; ***p<0.001; ns, not significant.

With respect to tau pathology relationships (assessed via temporal meta-ROI SUVR on tau-PET), IJV biomarkers showed nonsignificant trends towards stronger associations than their MCV counterparts did (**Fig. 3C, Fig. S5C**). Similarly, in the correlation with CSF biomarkers, Aβ42/40, p-tau217, p-tau217/Aβ42 and p-tau181/Aβ42 in IJV had steeper linear slopes than those in MCV (p<0.05) **(Fig. S7)**.

To quantify biomarker variance explained by AD pathologies, we found that cerebral Aβ status accounted for 16.97%–32.72% of the between-subject variation in MCV-Aβ42/40, increasing to 32.60%–49.28% for IJV-Aβ42/40 (p<0.05). However, cerebral Aβ and tau pathologies explained comparable variance proportions of the variance in p-tau217 and p-tau181 across both venous compartments **(Fig. 3B and D, Fig. S5B and D, Table S1)**.

### Performance in detecting cerebral pathologies

In differentiating Aβ-PET status, both analytical platforms demonstrated that IJV-Aβ42/40 had consistent superiority over MCV-Aβ42/40 in diagnostic performance (Lumipulse: AUC 0.899 vs. 0.849; 95% CI of difference, 0.062 to 0.137; Simoa: AUC 0.848 vs. 0.768; 95% CI, 0.052 to 0.107); conversely, IJV- and MCV-derived p-tau217, p-tau181, and their ratios to Aβ42 exhibited comparable AUC values **(Fig. 4A and E, eTables 1 and 2)**. Notably, Lumipulse IJV-Aβ42/40 achieved diagnostic performance (overall accuracy, sensitivity, specificity, PPV and NPV) comparable to those of the MCV-p-tau217 and MCV-p-tau217/Aβ42 measures **(Fig. 4C and G, eTable 4)**, with validation cohort replication confirming these findings **(Fig. 4I-L, eTables 3 and 5)**.

**Figure 4.**
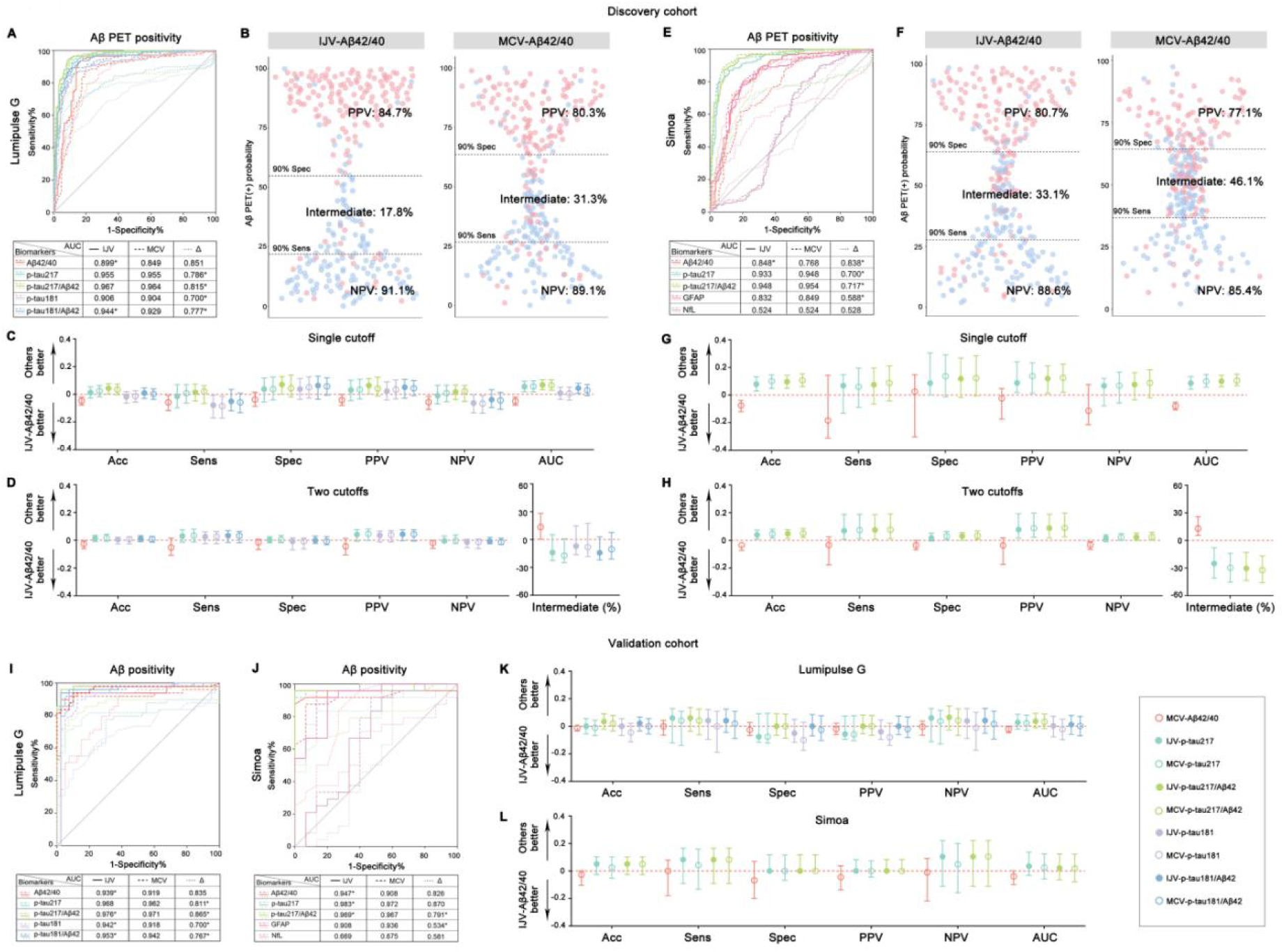
Performance of the IJV and MCV biomarkers for detecting cerebral Aβ pathology. (A,E,I,J) ROC curves and AUC values of the IJV and MCV biomarkers and the ΔC_IJV-MCV_ for Aβ PET positivity using the single-cutoff approach. (B,F) Diagnostic performance of Aβ42/40 in IJV and MCV in the classification of Aβ PET status using the two-cutoff approach. The upper and lower cutoffs were determined using 90% sensitivity/specificity. (C,D,G,H,K,L) Comparisons of the diagnostic metrics of IJV-Aβ42/40 with those of p-tau. (A-H) Performance of biomarkers for identifying Aβ PET positivity in the discovery cohort. (I-L) Performance of biomarkers for identifying cerebral Aβ status determined by Aβ PET or CSF Aβ42/40 in the validation cohort. (A-D,I,K) Biomarker assays on the Lumipulse G platform; (E-H,J,L) Biomarker assays using Simoa. Acc, accuracy; IJV, internal jugular vein; MCV, median cubital vein; NPV, negative predictive value; PET, positron emission tomography; PPV, positive predictive value; Sens, sensitivity; Spec, specificity. *Significantly different from the MCV biomarker.

Using a two-cutoff approach defined by 90% sensitivity/specificity, IJV-Aβ42/40 demonstrated enhanced accuracy (Lumipulse: 88.5% vs. 85.8%; 95% CI of difference, 0.3% to 6.1%; Simoa: 85.4% vs. 81.9%; 95% CI, 1.5% to 7.3%) and reduced indeterminate classification (Lumipulse: 17.8% vs. 31.3%; 95% CI, −28.3% to −0.3%; Simoa: 33.1% vs. 46.1%; 95% CI, −26.0% to −5.5%) compared with MCV-Aβ42/40 (**Fig. 4B and F**, **eTables 1 and 2**). Lumipulse IJV-Aβ42/40 maintained equivalent specificity and NPV to the MCV-p-tau217 biomarkers despite higher intermediate classification rates **(Fig. 4D and 4H, eTable 4**). These patterns persisted in sensitivity analyses using 95% sensitivity/specificity thresholds **(eTables 1, 2 and 4**).

For tau PET status classification, IJV-Aβ42/40 outperformed MCV-Aβ42/40 but remained inferior to MCV-p-tau217 **(Fig. S8, eTables 1 and 2)**.

In Aβ-positive patients with early tau pathology stage (tau PET Braak stage I-IV), compared with MCV-p-tau217, IJV-Aβ42/40 resulted in fewer false-negatives (1 out of 30 vs. 5 out of 30 misclassification) using a single cutoff, and fewer misclassification into definite negatives (2 vs. 4) and indeterminate cases (11 vs. 19) using two-cutoffs **(Table S2)**. While statistical significance was insignificant (p>0.05, likely due to the limited sample size), these trends suggest that IJV-Aβ42/40 may enhance early AD detection sensitivity compared with MCV-p-tau217.

## Discussion

The systemic metabolism of Aβ beyond the brain significantly contributes to the weak correlation between blood biomarkers and cerebral Aβ pathology, limiting its diagnostic utility. Brain-derived Aβ crosses the blood-brain barrier (BBB) into the circulation and exists in free form or is bound to carriers (e.g., albumin) before being cleared via peripheral organs (i.e., the kidney and liver) and cells^16–19^. Additionally, blood Aβ originates not only from the brain but also from peripheral sources (i.e., platelets and red blood cells)^20,21^. These dual pathways necessitate strategies to minimize peripheral interference to improve the diagnostic performance of blood biomarkers^22^.

To amplify intergroup fold changes and mitigate peripheral metabolic effects, we implemented IJV sampling. Proximally located to the brain and clinically accessible, IJV blood contains brain-derived proteins prior to peripheral metabolism. This approach mirrors established endocrine diagnostic methods (e.g., adrenal venous sampling for primary aldosteronism^23,24^; parathyroid venous sampling for hyperparathyroidism^25^). In the RA subcohort, we observed an increase of ∼8.5% to 12.5% in Aβ42 levels and ∼14.8% to 15.2% in Aβ40 levels in the IJV compared to the RA (data not shown) and similar levels of Aβ42 and Aβ40 between the RA and the MCV. These results align with a previous study, which reported an ∼7.5% increase in Aβ concentration in the cerebral vein (encompassing the sigmoid sinus, inferior petrosal sinus, and the internal jugular vein) relative to the RA, as well as comparable Aβ levels between the RA and the peripheral vein^26^. In our study, IJV-Aβ42/40 demonstrated superior performance: a ∼25% reduction in AD patients (vs. ∼15% for MCV-Aβ42/40), a stronger correlation with cerebral Aβ burden, and enhanced diagnostic accuracy. Crucially, IJV-Aβ42/40 correlated with Aβ PET Centiloid severity in Aβ(+) populations, addressing a gap where blood Aβ levels failed to reflect brain deposition magnitude.

Given comparable biomarker levels in the MCV and RA, we selected ΔC_IJV-MCV_ over ΔC_IJV-RA_ for clinical practicality to reflect the net efflux of brain-derived proteins into systemic circulation and their subsequent clearance in the periphery. In our study, we observed marked reductions in %ΔAβ42 in Aβ(+) individuals, mirroring CSF trends in AD patients. A previous study also reported that the IJV-Aβ levels, rather than peripheral plasma Aβ, had a parallel change with CSF Aβ changes after IVIg administration^27^. Biomarker ratios (%ΔAβ42/40, %Δp-tau/Aβ42) showed greater resistance to peripheral interference than individual biomarkers did. For example, cerebral Aβ status explained only 16.4% of MCV-Aβ42 variability but 32.7% of MCV-Aβ42/40 variability (Lumipulse G). This aligns with reports that CKD elevates individual Aβ isoforms but not Aβ42/40 ^8^, likely due to shared peripheral clearance pathways across isoforms. Combining IJV sampling with ratio-based biomarkers further reduced systemic noise, increasing cerebral Aβ-driven variability to 49.3% (vs. 32.7% for MCV-Aβ42/40).

Furthermore, compared with MCV sampling, IJV sampling increased the diagnostic accuracy of Aβ42/40 for cerebral amyloid pathology from 72.4%-88.8% to 79.9%-92.9%, while reducing indeterminate classifications by 28%-43% using a two-cutoff approach. Although plasma p-tau217 has exhibited excellent performance in detecting cerebral amyloid pathology^3,4^, IJV-Aβ42/40 demonstrated equivalent clinical accuracy to both p-tau217 and p-tau217/Aβ42 ratio, although the intermediate percentage for IJV-Aβ42/40 was greater. Notably, IJV-Aβ42/40 showed superior performance trends in early-stage patients compared with MCV-p-tau217, with higher accuracy and fewer intermediate results. Collectively, our findings establish IJV-Aβ42/40 as an accurate and potentially early biomarker for cerebral amyloid pathology.

The reliability of our results lies in cross-validation across two independent cohorts using dual biomarker detection platforms. Our study has several limitations. (1) Early-stage cohort size: The small early-stage sample limited analysis to misclassification rates, necessitating validation in large-scale early-stage AD cohorts. (2) Tauopathy representation: Rare non-AD tauopathies with extensive tau pathology (e.g., NIID, CJD, corticobasal degeneration [CBD]) were underrepresented, where p-tau elevations may confound the diagnosis and IJV-Aβ42/40 could demonstrate AD specificity. (3) Comorbidity data: Few participants had significant systemic abnormalities such as CKD, precluding assessment of the contribution of peripheral metabolism to biomarker variability. Future studies should identify confounding factors in heterogeneous populations with high comorbidity burdens and evaluate whether IJV-Aβ42/40 measurements mitigate these effects.

In summary, IJV sampling establishes blood Aβ42/40 as a severity-sensitive biomarker for cerebral amyloidosis, achieving p-tau-level diagnostic accuracy. By reducing peripheral metabolic interference, this strategy offers a novel framework for optimizing blood-based AD diagnostics.

## Supporting information

eMethods; eResults; Fig. S1; Fig. S2; Fig. S3; Fig. S4; Fig. S5; Fig. S6; Fig. S7; Fig. S8; Table S1; Table S2

## Data Availability

All data produced in the present study are available upon reasonable request to the corresponding author. The request will be reviewed by the CADS and TBRAIN investigators and relevant ethics boards to verify whether the request is subject to any intellectual property or confidentiality obligations. A data sharing agreement must be obtained prior to release.

## Author contributions

YJ Wang and J Wang: study design; J Wang, S Huang and N Wu: data analysis; DY Fan, C Liu, PW Zhao, XL Gao, QZ Wang, Y Li, B Liu, YY Ma, RC Zhao, YP Zhu, QY Li, XY Liu, X Chen, YJ Lai, F Zeng, YH Liu, XL Bu: sample and data collection; J Wang and YJ Wang manuscript drafting; T Guo, Y Tang, JT Yu, CL Masters, J Guo, J Yang, Q Mao: critical revision for important intellectual content. All authors reviewed and approved the final version of the manuscript.

## Acknowledgements

The study was supported by the National Key Research and Development Program of China (2023YFC3605400 to Y.-J. W.), Joint project of the Chongqing Science and Technology Bureau and the Health Commission (2024GGXM003 to Y.-J.W.). We thank all the research volunteers who participated in CADS and TBRAIN studies from which these data were obtained and their supportive families and caregivers.

## Data sharing

Raw and analyzed de-identified data can be requested from the corresponding author. The request will be reviewed by the CADS and TBRAIN investigators and relevant ethics boards to verify whether the request is subject to any intellectual property or confidentiality obligations. A data sharing agreement must be obtained prior to release.

## References

1. Dubois B, Villain N, Schneider L, et al. Alzheimer Disease as a Clinical-Biological Construct—An International Working Group Recommendation. JAMA Neurol. Published online November 1, 2024. doi:10.1001/jamaneurol.2024.3770

2. Jack CR, Andrews JS, Beach TG, et al. Revised criteria for diagnosis and staging of Alzheimer’s disease: Alzheimer’s Association Workgroup. Alzheimers Dement. Published online June 27, 2024:alz.13859. doi:10.1002/alz.13859

3. Barthélemy NR, Salvadó G, Schindler SE, et al. Highly accurate blood test for Alzheimer’s disease is similar or superior to clinical cerebrospinal fluid tests. Nat Med. Published online February 21, 2024. doi:10.1038/s41591-024-02869-z

4. Ashton NJ, Brum WS, Di Molfetta G, et al. Diagnostic Accuracy of a Plasma Phosphorylated Tau 217 Immunoassay for Alzheimer Disease Pathology. JAMA Neurol. Published online January 22, 2024. doi:10.1001/jamaneurol.2023.5319

5. Kurihara M, Komatsu H, Sengoku R, et al. CSF P-Tau181 and Other Biomarkers in Patients With Neuronal Intranuclear Inclusion Disease. Neurology. 2023;100(10):e1009–e1019. doi:doi: 10.1212/WNL.0000000000201647

6. Emeršič A, Ashton NJ, Vrillon A, et al. Cerebrospinal fluid p-tau181, 217, and 231 in definite Creutzfeldt–Jakob disease with and without concomitant pathologies. Alzheimers Dement. 2024;20(8):5324–5337. doi:doi: 10.1002/alz.13907

7. Mammel AE, Hsiung GR, Mousavi A, et al. Clinical decision points for two plasma p-tau217 laboratory developed tests in neuropathology confirmed samples. Alzheimers Dement Diagn Assess Dis Monit. 2025;17(1):e70070. doi:10.1002/dad2.70070

8. Syrjanen JA, Campbell MR, Algeciras-Schimnich A, et al. Associations of amyloid and neurodegeneration plasma biomarkers with comorbidities. Alzheimers Dement. 2022;18(6):1128–1140. doi:10.1002/alz.12466

9. Mielke MM, Dage JL, Frank RD, et al. Performance of plasma phosphorylated tau 181 and 217 in the community. Nat Med. 2022;28(7):1398–1405. doi:10.1038/s41591-022-01822-2

10. Wang JH, Wu YJ, Tee BL, Lo RY. Medical Comorbidity in Alzheimer’s Disease: A Nested Case-Control Study. Fink A, ed. J Alzheimers Dis. 2018;63(2):773–781. doi:10.3233/JAD-170786

11. Valletta M, Vetrano DL, Rizzuto D, et al. Blood biomarkers of Alzheimer’s disease in the community: Variation by chronic diseases and inflammatory status. Alzheimers Dement. 2024;20(6):4115–4125. doi:10.1002/alz.13860

12. Wang J, Fan DY, Li HY, et al. Dynamic changes of CSF sPDGFRβ during ageing and AD progression and associations with CSF ATN biomarkers. Mol Neurodegener. 2022;17(1):9. doi:10.1186/s13024-021-00512-w

13. Fan DY, Jian JM, Huang S, et al. Establishment of combined diagnostic models of Alzheimer’s disease in a Chinese cohort: the Chongqing Ageing & Dementia Study (CADS). Transl Psychiatry. 2022;12(1):252. doi:10.1038/s41398-022-02016-7

14. Seibyl JP, DuBois JM, Racine A, et al. A Visual Interpretation Algorithm for Assessing Brain Tauopathy with ^18^ F-MK-6240 PET. J Nucl Med. 2023;64(3):444–451. doi:10.2967/jnumed.122.264371

15. Bourgeat P, Doré V, Fripp J, et al. Implementing the centiloid transformation for 11C-PiB and β-amyloid 18F-PET tracers using CapAIBL. NeuroImage. 2018;183:387–393. doi:10.1016/j.neuroimage.2018.08.044

16. Wang J, Gu BJ, Masters CL, Wang YJ. A systemic view of Alzheimer disease — insights from amyloid-β metabolism beyond the brain. Nat Rev Neurol. 2017;13(10):612–623. doi:10.1038/nrneurol.2017.111

17. Xiang Y, Bu XL, Liu YH, et al. Physiological amyloid-beta clearance in the periphery and its therapeutic potential for Alzheimer’s disease. Acta Neuropathol (Berl). 2015;130(4):487–499. doi:10.1007/s00401-015-1477-1

18. Tian DY, Cheng Y, Zhuang ZQ, et al. Physiological clearance of amyloid-beta by the kidney and its therapeutic potential for Alzheimer’s disease. Mol Psychiatry. 2021;26(10):6074–6082. doi:10.1038/s41380-021-01073-6

19. Cheng Y, He CY, Tian DY, et al. Physiological β-amyloid clearance by the liver and its therapeutic potential for Alzheimer’s disease. Acta Neuropathol (Berl). 2023;145(6):717–731. doi:10.1007/s00401-023-02559-z

20. Sun HL, Chen SH, Yu ZY, et al. Blood cell-produced amyloid-β induces cerebral Alzheimer-type pathologies and behavioral deficits. Mol Psychiatry. 2021;26(10):5568–5577. doi:10.1038/s41380-020-0842-1

21. Roher AE, Esh CL, Kokjohn TA, et al. Amyloid beta peptides in human plasma and tissues and their significance for Alzheimer’s disease. Alzheimers Dement. 2009;5(1):18–29. doi:10.1016/j.jalz.2008.10.004

22. Wang J, Chen M, Masters CL, Wang Y. Translating blood biomarkers into clinical practice for Alzheimer’s disease: Challenges and perspectives. Alzheimers Dement. 2023;19(9):4226–4236. doi:10.1002/alz.13116

23. Reincke M, Bancos I, Mulatero P, Scholl UI, Stowasser M, Williams TA. Diagnosis and treatment of primary aldosteronism. Lancet Diabetes Endocrinol. 2021;9(12):876–892. doi:10.1016/S2213-8587(21)00210-2

24. Liu C, Zheng F, Zhang X, Pan J, Ding W, Tian X. Selective venous sampling for secondary hypertension. Hypertens Res. 2024;47(7):1766–1778. doi:10.1038/s41440-024-01699-3

25. Zolin SJ, Crawford K, Rudin AV, et al. Selective parathyroid venous sampling in reoperative parathyroid surgery: A key localization tool when noninvasive tests are unrevealing. Surgery. 2021;169(1):126–132. doi:10.1016/j.surg.2020.05.014

26. Roberts KF, Elbert DL, Kasten TP, et al. Amyloid-β efflux from the central nervous system into the plasma. Ann Neurol. 2014;76(6):837–844. doi:10.1002/ana.24270

27. Kasai T, Kondo M, Ishii R, et al. Aβ levels in the jugular vein and high molecular weight Aβ oligomer levels in CSF can be used as biomarkers to indicate the anti-amyloid effect of IVIg for Alzheimer’s disease. Valera E, ed. PLOS ONE. 2017;12(4):e0174630. doi:10.1371/journal.pone.0174630

