## Supplementary material for "Performance of blood biomarkers in internal jugular vein for Alzheimer disease pathologies: the Delta Study": eMethods; eResults; Fig. S1; Fig. S2; Fig. S3; Fig. S4; Fig. S5; Fig. S6; Fig. S7; Fig. S8; Table S1; Table S2

**eTable 1**. Performance of the IJV and MCV biomarkers measured on the Lumipulse G platform in the classification of Aβ and tau statuses in the discovery cohort.

**eTable 2.** Performance of the IJV and MCV biomarkers measured on the Simoa platform in the classification of Aβ and tau statuses in the discovery cohort.

**eTable 3.** Performance of the IJV and MCV biomarkers in the classification of Aβ status in the validation cohort.

**eTable 4.** Comparisons in performance of IJV-Aβ42/40 and p-tau for identifying Aβ PET positivity in the discovery cohort.

**eTable 5.** Comparisons in performance of IJV-Aβ42/40 and p-tau for identifying Aβ PET positivity in the validation cohort.

### eMethods

#### Inclusion and exclusion criteria

Patients must meet all the following criteria: 1) had a complaint of cognitive decline or consult for the risk of AD at the memory clinic at three hospitals, and Alzheimer's disease (AD) was the possible etiology or need to be excluded for the etiology dignosis by the dementia specialist; 2) was willing to participate in this study and completed blood sampling from the internal jugular vein (IJV) and median cubital vein (MCV); 3) complete Aβ-PET scan or CSF sampling; 4) the patient might benefit from the knowledge of brain amyloid status by the dementia specialist; 5) the patient wanted to know the brain amyloid status.

The exclusion criteria includes: 1) had an unstable medical or psychiatric disease, or other conditions that could interfere with the completion of an Aβ-PET scan or CSF or blood sampling; 2) refuse to participate in this study.

#### Clinical assessments

Participants underwent clinical assessments, including physical examination, laboratory tests, APOE genotyping, magnetic resonance imaging (MRI), neuropsychological tests (including Minimum Mental State Examination [MMSE], Montreal Cognitive Assessment [MoCA], Clinical Dementia Rating [CDR], Activities of Daily Living [ADL], etc.), and PET scans. Demographic information (age, sex and years of education), medical history, comorbidities (including diabetes, hypertension, dislipidemia, history of stroke or myocardial infarction [MI], atrial fibrillation, cancer, etc.) and medication use were collected.

#### Blood and CSF sampling and processing

Three centers adopted the same standard operating procedure (SOP) of biofluid collection and processing. Fasting blood from IJV and MCV were collected into

EDTA-containing tubes using butterfly needles of the same size within 1 hour. In the radial artery (RA) subcohort of CADS, RA blood was also collected within 1 hour using the same blood collection device. IJV blood sampling was completed by experienced anesthesiologists under the guidence of ultrasound, and MCV and RA blood was collected by experienced nurses. In the validation cohort, only IJV and MCV blood were collected.

Blood was left to stand for half an hour at room temperature and then centrifuged at 2000 × *g* for 10 min at room temperature to separate the plasma. The plasma samples were aliquoted and stored at -80 °C within 2 hours of the blood collection.

CSF samples were collected via lumbar puncture by experienced doctors at the same day. Within 2 hours, samples were centrifuged (2000 × g for 10 min at room temperature), aliquoted, and stored at -80 °C.

**Measurements of blood and CSF biomarkers**

Plasma biomarkers were measured at Daping hospital: (1) Lumipulse plasma Aβ42, Aβ40, p-tau181, and p-tau217 were measured using commercially available kits (Fujirebio Europe, Ghent, Belgium); (2) Simoa plasma Aβ42, Aβ40, GFAP, NfL were measured using commercial kit Human Neurology 4-Plex E kit and Simoa p-tau217 were measured using ALZpath p-tau217 kit on HD-X analyzer (Fujirebio Europe, Ghent, Belgium).

**PET acquisition and analysis**

Aβ PET was evaluated by visual read according to an FDA-approved protocol, and tau PET was visually analysed using a dichotomous negative/positive tau PET read according to the visual read algorithm used by Seibyl et al for assessing $[^{18}F]$MK-6240 PET scans by two experienced nuclear medicine physicians. Meanwihle, both Aβ and tau PET images were analysed using Computational

Analysis of PET by AIBL (CapAIBL), a publicly available cloud-based platform in which PET images are spatially normalized to a standard template via an adaptive atlas approach (https://capaibl-milxcloud.csiro.au). The Aβ PET scans were quantified using standard centiloids (CLs), and the tau PET scans were quantified using the meta- temporal standard uptake value ratio (SUVR).

**Statistical analysis**

The normal distribution of continuous data was assessed by the Shapiro-Wilk test and by visually inspecting the Q-Q plot. Continuous variables were shown as means (standard deviations [SD]) or median (interquartile range [IQR]). Comparisons on biomarmer levels between IJV and MCV were assessed by the paired t-test or Wilcoxon matched-pairs signed rank test, as appropriate. Between-group differences in %Δ of biomarkers were evaluated by the Mann-Whitney U test or the Kruskal-Wallis test. Categorical variables were expressed as counts and percentages and compared using the Chi-squared test. Correlations were analyzed using the Pearson r or the Spearman ρ as appropriate, and the correlation coefficients were compared using the Fisher Z-transformation. The slopes of the two regression lines were compared using the F-test. Univariate linear regression models with the coefficient of determination ($R^2$) were used to quantify the contributions of brain pathologies to blood biomarkers.

To evaluate the diagnostic performance of IJV and MCV biomarkers, the single-cutoff approach derives a binary reference (positive/negative) based on the max Youden index using the receiver operating characteristic (ROC) curves (cutpointr package). The two-cutoffs approach derives the three-range references (positive/uncertain/negative). The lower threshold was obtained by maximizing the specificity with the sensitivity fixed at 90% and 95% respectively, whereas the upper threshold was obtained by maximizing the sensitivity with the specificity fixed at 90% and 95%. Participants with biomarker levels between these two thresholds were categorized as intermediate. The differences in diagnostic metrics (including accuracy and intermediate percentage between IJV and MCV biomarkers were calculated as the mean with 95% CI of the bootstrapped sample (n = 1,000 resamples with replacement stratifying by the output), and were considered equivalent if the 95% CI for the mean difference included zero.

### eResults

#### Comparisons of blood biomarker levels between the MCV and RA

First, we compared biomarker levels between the MCV and RA in the RA subcohort and found strong correlations and similar levels of biomarkers between the two sites. The results revealed strong correlations of biomarker levels between the two sites. No significant differences in AD core biomarker levels between the RA and MCV were detected by Simoa assays ($p>0.05$). The Lumipulse G assays showed that the levels of AD core biomarkers in RA were lower than those in MCV, but the differences ($\%\Delta = [C_{RA}-C_{MCV}]/C_{MCV}\times100\%$) were tiny (median %ΔAβ42: -1.46%; %ΔAβ40: -1.06%; %Δp-tau217: -4.76%; %Δp-tau181: -6.67%) (**Fig. S2**). These findings suggest that AD biomarker levels in MCV were similar to those of RA and that differences in biomarker levels between IJV and MCV, to some extent, may reflect the amount of brain-derived molecules flowing from the brain to the circulation.

**Figure S1. Correlations of blood biomarker levels measured on the Lumipulse G and Simoa platforms.**

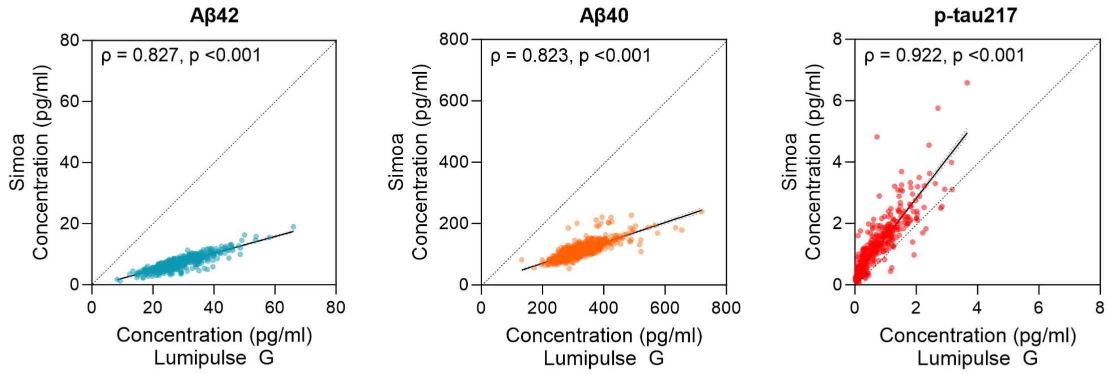

**Figure S2. Correlations of blood biomarker levels between the MCV and IJV.**

IJV, internal jugular vein; MCV, median cubital vein. ***p<0.001.

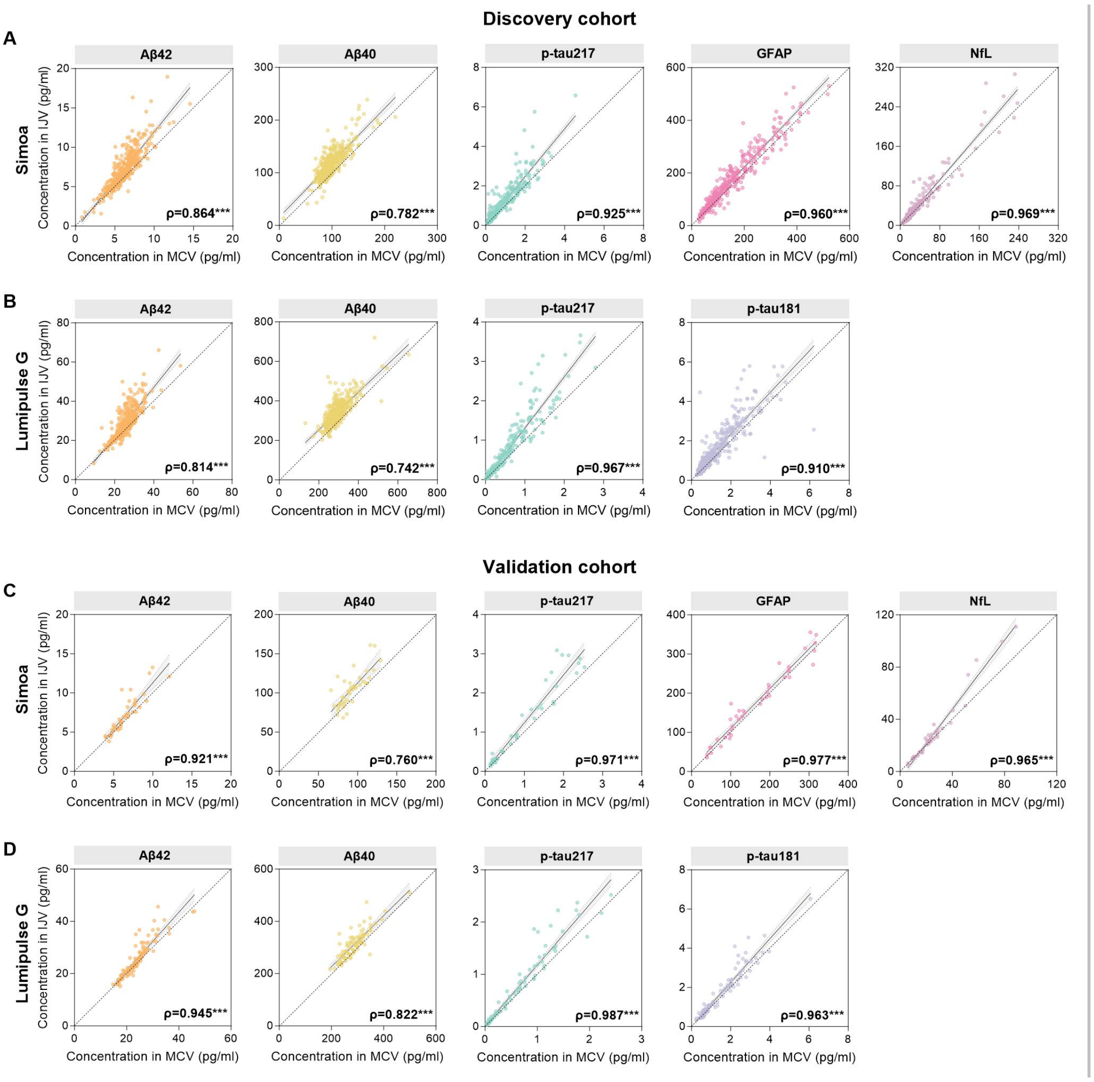

**Figure S3. Comparisons of blood biomarker levels between the MCV and RA in the RA subcohort.** (A,C) Correlations of biomarker levels between the MCV and the RA. (B,D) Comparisons of blood biomarker levels between the MCV and the RA and the differences between them in the entire cohort. $\%\Delta C_{RA\text{-}MCV}$ of biomarkers were calculated as: $(C_{RA}\text{-}C_{MCV})/C_{MCV}*100\%$. Comparisons of biomarker levels between MCV and IJV were analyzed using paired t-test for Aβ42, Aβ40 and Aβ42/40, and using Wilcoxon matched-pairs signed rank test for others. MCV, median cubital vein; RA, radial artery. $C_{RA}$, concentration in RA; $C_{MCV}$, concentration in MCV. *p<0.05; **p<0.01; ***p<0.001; ns, not significant.

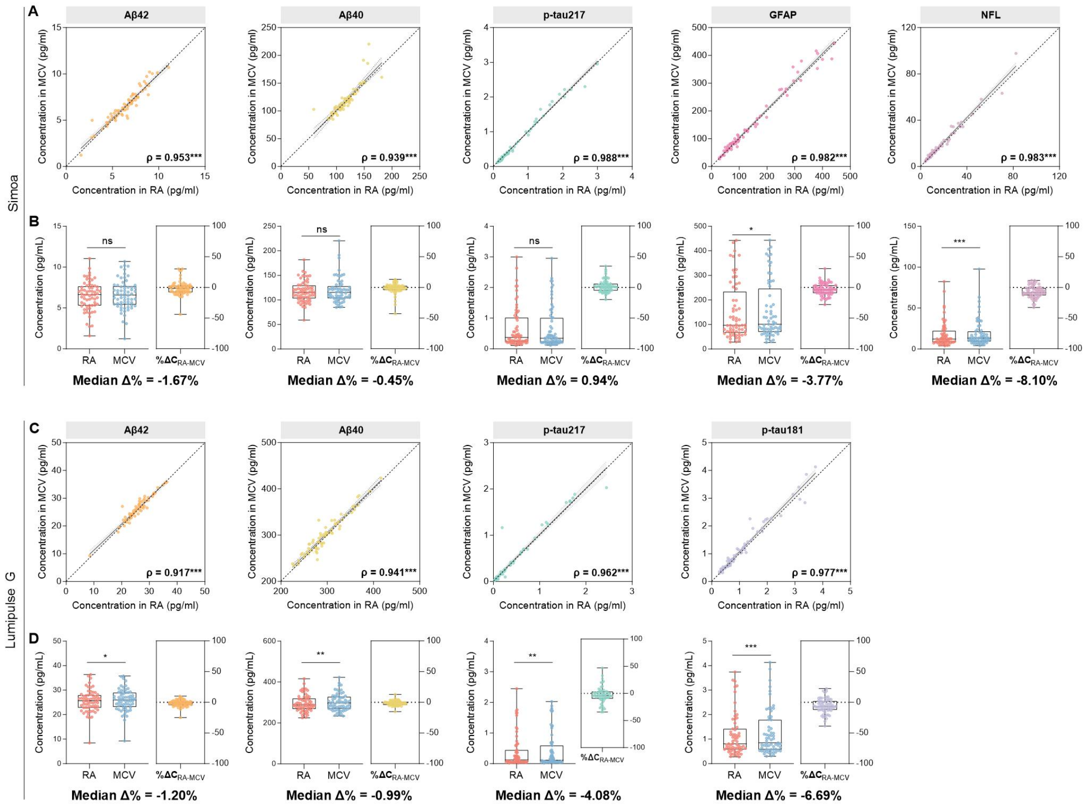

**Figure S4. Comparisons of blood biomarker levels between the MCV and IJV in the validation cohort. (A,C)** The levels of AD biomarkers in the IJV and MCV. **(B,D)** The differences in blood biomarker levels between the MCV and IJV in the entire cohort and Aβ-positive and Aβ-negative subgroups. $\%\Delta C_{IJV\text{-}MCV}$ of biomarkers was calculated as: $(C_{IJV}-C_{MCV})/C_{MCV}*100\%$. Aβ-positive (Aβ+) or Aβ-negative (Aβ-) status was defined by Aβ PET or CSF Aβ42/40 ratio. (A,B) Biomarkers were measured using Simoa methods (n=40); (C,D) Biomarkers were measured using Lumipulse G methods (n=92). IJV, internal jugular vein; MCV, median cubital vein. $C_{IJV}$, concentration in the IJV; $C_{MCV}$, concentration in the MCV. Comparisons of biomarker levels between the MCV and IJV were performed using paired t-test for Aβ42, Aβ40 and Aβ42/40, and using Wilcoxon matched-pairs signed rank test for others. *p<0.05; **p<0.01; ***p<0.001; ns, not significant.

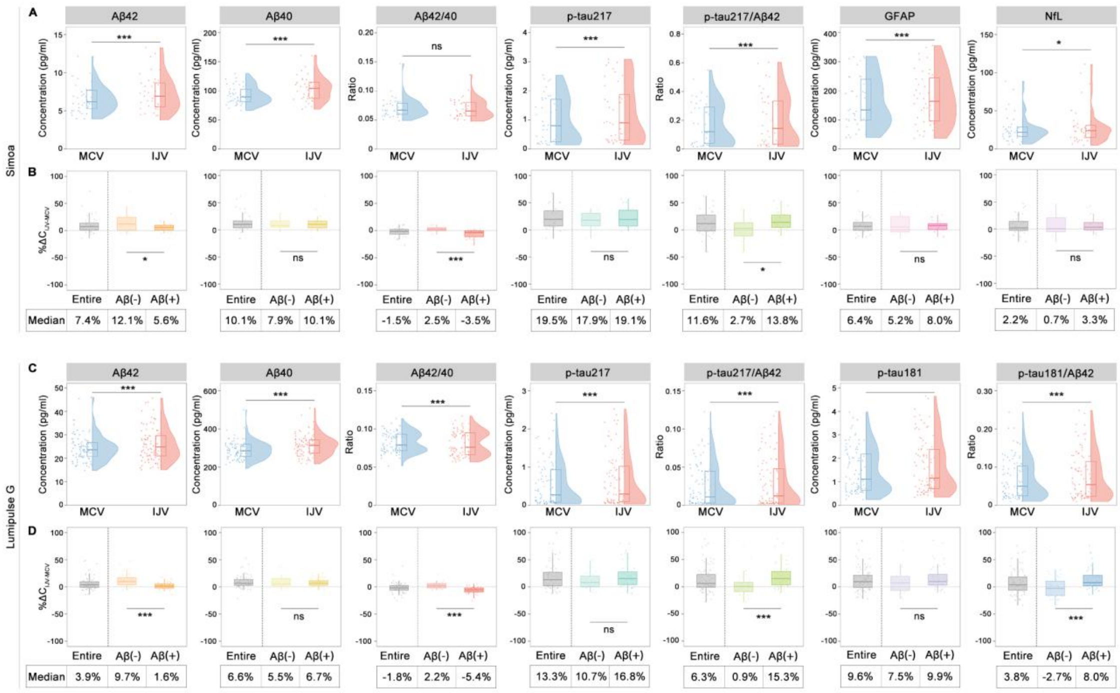

**Figure S5. Correlations of the IJV and MCV biomarkers measured by Simoa with brain burden of Aβ and tau pathologies. (A)** Correlations of the IJV and MCV biomarkers with brain Aβ burden measured by Aβ-PET Centiloids. **(B)** Contribution of brain Aβ status to the IJV and MCV biomarkers. Aβ status was defined by visual reading of Aβ PET imaging or CSF Aβ42/40 ratio. **(C)** Correlations of the IJV and MCV biomarkers with brain tau pathology measured by tau-PET meta-temporal SUVR. **(D)** Contribution of brain tau status to the IJV and MCV biomarkers. Tau status was defined by visual reading of Tau PET imaging. CSF, cerebrospinal fluid; IJV, internal jugular vein; MCV, median cubital vein; PET, positron emission tomography; SUVR, standardized uptake value ratio. $p_\rho$, p value for Spearman correlation coefficient ρ; $p_{slope}$, p value for slopes of the fitted linear. *p<0.05; **p<0.01; ***p<0.001; ns, not significant.

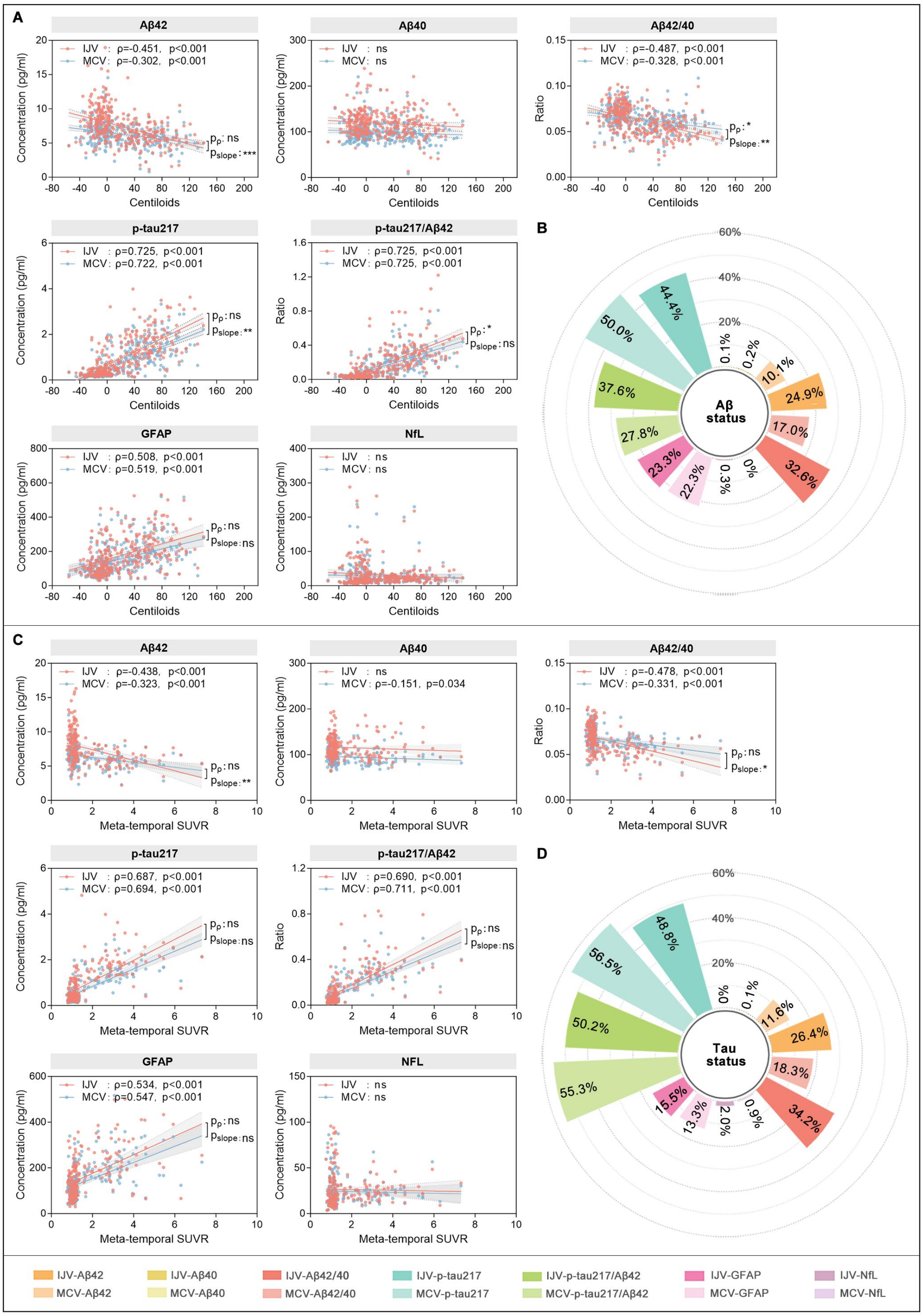
A
Aβ42
Aβ40
Aβ42/40
p-tau217
p-tau217/Aβ42
GFAP
NfL
Centiloids
Concentration (pg/ml)
Ratio
B
Aβ status
C
NFL
Meta-temporal SUVR
D
Tau status
IJV-Aβ42
MCV-Aβ42
IJV-Aβ40
MCV-Aβ40
IJV-Aβ42/40
MCV-Aβ42/40
IJV-p-tau217
MCV-p-tau217
IJV-p-tau217/Aβ42
MCV-p-tau217/Aβ42
IJV-GFAP
MCV-GFAP
IJV-NfL
MCV-NfL

**Figure S6. Correlations of the IJV and MCV biomarkers and $\Delta C_{IJV\text{-}MCV}$ with Aβ PET Ccentiloids in Aβ-positive populations.** Aβ-positive was defined by Centiloid >15. IJV, internal jugular vein; MCV, median cubital vein; PET, positron emission tomography. ns, not significant.

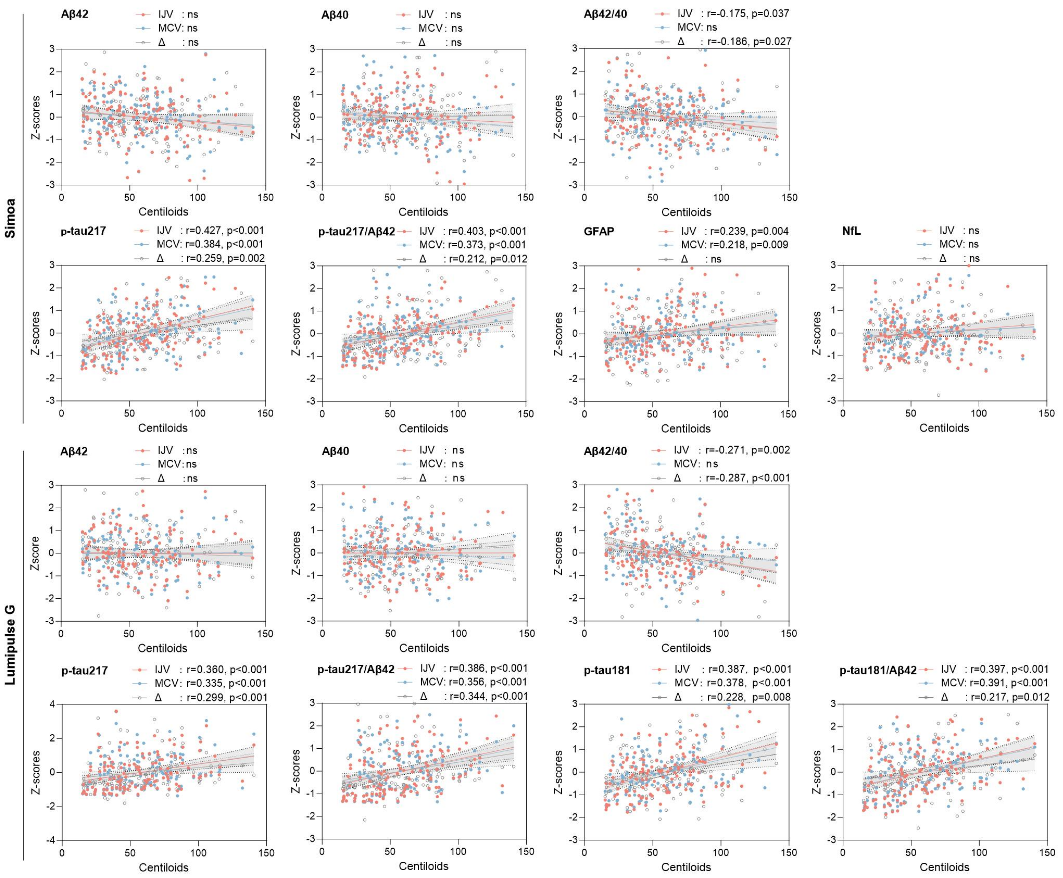

**Figure S7. Correlations of the IJV and MCV biomarkers measured by Lumipulse G with CSF biomarkers.** CSF, cerebrospinal fluid; IJV, internal jugular vein; MCV, medial cubital vein. $p_\rho$, p value for Spearman correlation coefficient ρ; $p_{slope}$, p value for slopes of the fitted linear. *p<0.05; **p<0.01; ***p<0.001; ns, not significant.

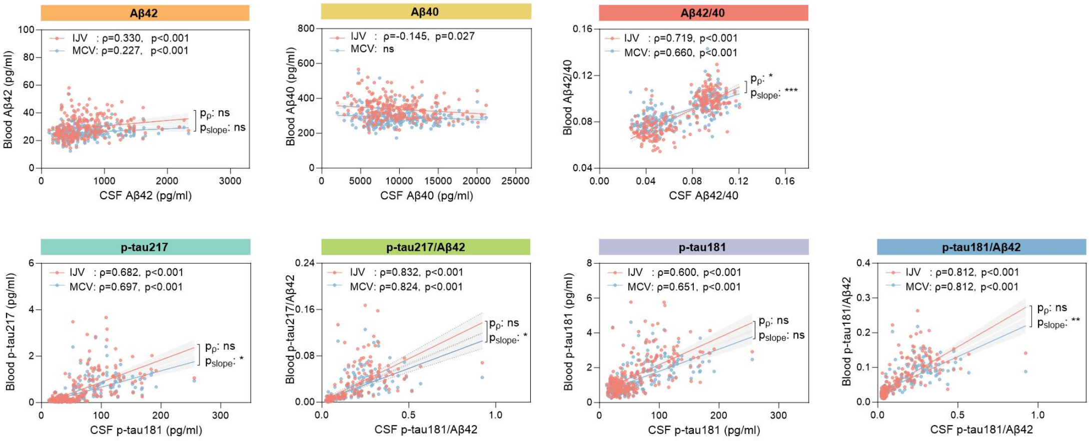

**Figure S8. Performance of the IJV and MCV biomarkers in identifying tau PET positivity in the discovery cohort.** (A, B) ROC curves and AUC values of the IJV and MCV biomarkers and the $\Delta C_{IJV\text{-}MCV}$ using the single-cutoff approach. (C,D) Diagnostic metrics of the IJV and MCV biomarkers using the two-cutoff approach. The upper and lower cutoffs were determined using 95% sensitivity/specificity. (E) Comparisons in diagnostic metrics of IJV-Aβ42/40 with p-tau. (A,C) Biomarker assays on the Lumipulse G platform; (B,D) Biomarker assays using Simoa. Acc, accuracy; IJV, internal jugular vein; MCV, median cubital vein; NPV, negative predictive value; PET, positron emission tomography; PPV, positive predictive value; Sens, sensitivity; Spec, specificity.

*Significantly different from MCV biomarker.

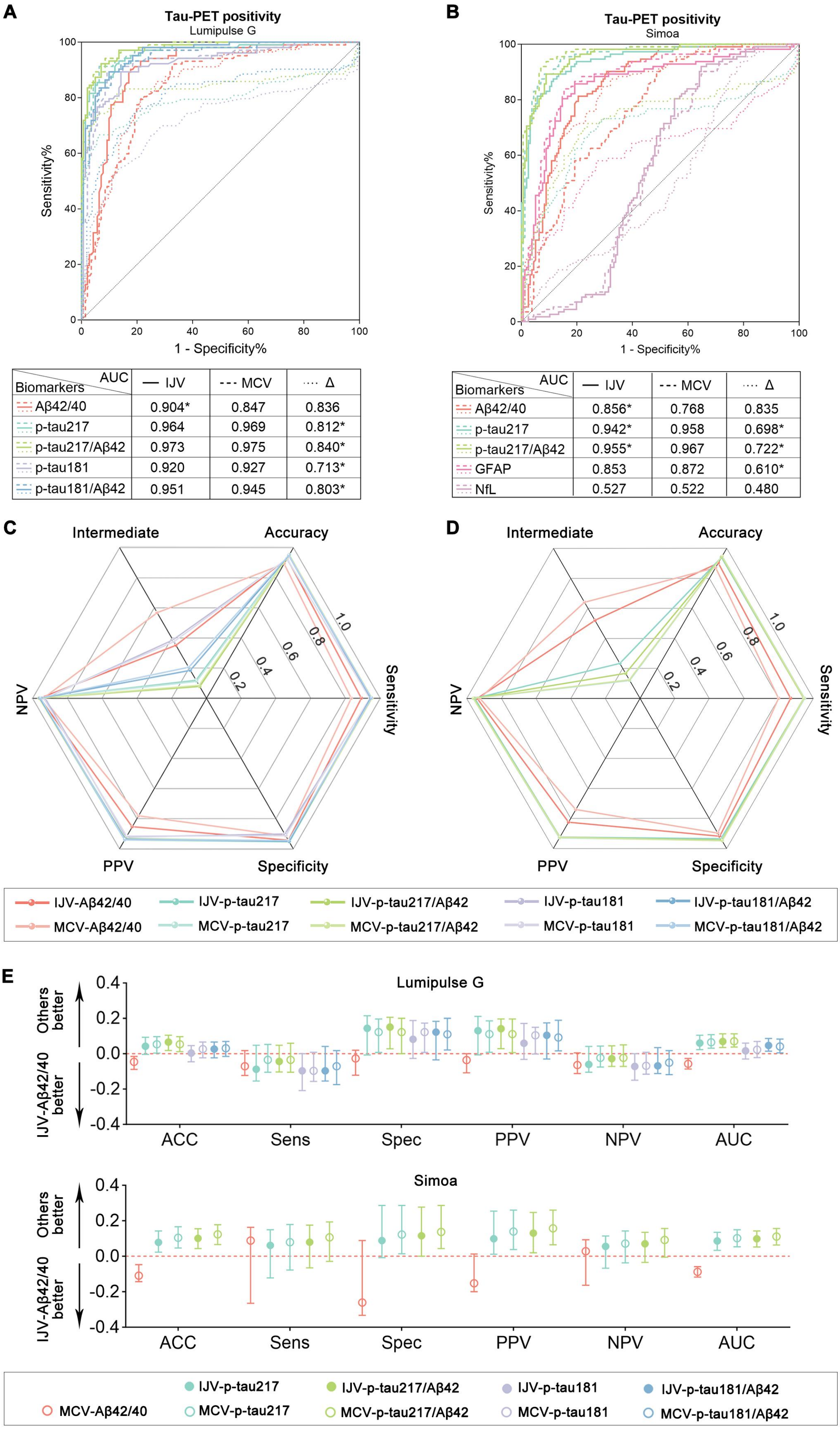

| Biomarkers \ AUC | — IJV | --- MCV | ···· Δ |
|---|---|---|---|
| Aβ42/40 | 0.904* | 0.847 | 0.836 |
| p-tau217 | 0.964 | 0.969 | 0.812* |
| p-tau217/Aβ42 | 0.973 | 0.975 | 0.840* |
| p-tau181 | 0.920 | 0.927 | 0.713* |
| p-tau181/Aβ42 | 0.951 | 0.945 | 0.803* |

| Biomarkers \ AUC | — IJV | --- MCV | ···· Δ |
|---|---|---|---|
| Aβ42/40 | 0.856* | 0.768 | 0.835 |
| p-tau217 | 0.942* | 0.958 | 0.698* |
| p-tau217/Aβ42 | 0.955* | 0.967 | 0.722* |
| GFAP | 0.853 | 0.872 | 0.610* |
| NfL | 0.527 | 0.522 | 0.480 |

**Table S1. Contributions of brain Aβ and tau statuses to between-subject variations in the IJV and MCV biomarkers.**

| Simoa | | | | Lumipulse G | | | |
|---|---|---|---|---|---|---|---|
| | IJV | MCV | P value | | IJV | MCV | P value |
| explained by brain Aβ status | | | | explained by Aβ status | | | |
| Aβ42 | 24.90% | 10.05% | **0.004** | Aβ42 | 31.25% | 16.40% | **0.012** |
| Aβ40 | 0.21% | 0.11% | 0.301 | Aβ40 | 0.11% | 0.20% | 0.354 |
| Aβ42/40 | 32.60% | 16.97% | **0.006** | Aβ42/40 | 49.28% | 32.72% | **0.004** |
| p-tau217 | 44.36% | 49.98% | 0.314 | p-tau217 | 39.69% | 42.38% | 0.651 |
| p-tau217/Aβ42 | 37.58% | 27.77% | 0.094 | p-tau217/Aβ42 | 41.60% | 43.03% | 0.802 |
| GFAP | 23.33% | 22.37% | 0.864 | p-tau181 | 41.86% | 37.58% | 0.475 |
| NfL | 0.31% | 0% | 0.511 | p-tau181/Aβ42 | 52.13% | 48.86% | 0.548 |
| explained by Tau-PET status | | | | explained by Tau-PET status | | | |
| Aβ42 | 26.42% | 11.56% | **0.016** | Aβ42 | 31.25% | 14.98% | **0.016** |
| Aβ40 | 0.07% | 0.04% | 0.591 | Aβ40 | 0.06% | 0.27% | 0.443 |
| Aβ42/40 | 34.22% | 18.32% | **0.017** | Aβ42/40 | 48.16% | 32.60% | **0.018** |
| p-tau217 | 48.86% | 56.55% | 0.205 | p-tau217 | 49.84% | 53.29% | 0.582 |
| p-tau217/Aβ42 | 50.13% | 55.35% | 0.399 | p-tau217/Aβ42 | 50.98% | 55.06% | 0.505 |
| GFAP | 15.44% | 13.32% | 0.710 | p-tau181 | 48.58% | 49.00% | 0.937 |
| NfL | 1.99% | 0.95% | 0.617 | p-tau181/Aβ42 | 57.61% | 58.98% | 0.795 |

**Table S2. Accuracy of Aβ42/40 and other biomarkers in Aβ-positive patients with tau PET Braak stage I-IV.**

| Lumipulse G | | Single-cutoff | | | | Two-cutoffs | | | | | |
|---|---|---|---|---|---|---|---|---|---|---|---|
| | | Negative | | Positive | | Negative | | Intermediate | | Positive | |
| | | No. | % | No. | % | No. | % | No. | % | No. | % |
| Aβ42/40 | IJV | 1 | 3.33 | 29 | 96.67 | 2 | 6.67 | 11 | 36.67 | 17 | 56.67 |
| | MCV | 1 | 3.33 | 29 | 96.67 | 1 | 3.33 | 18 | 60.00 | 11 | 36.67 |
| p-tau217 | IJV | 6 | 20.00 | 24 | 80.00 | 3 | 10.00 | 12 | 40.00 | 15 | 50.00 |
| | MCV | 5 | 16.67 | 25 | 83.33 | 4 | 13.33 | 19 | 63.33 | 7 | 23.33 |
| p-tau181 | IJV | 8 | 26.67 | 22 | 73.33 | 4 | 13.33 | 14 | 46.67 | 12 | 40.00 |
| | MCV | 9 | 30.00 | 21 | 70.00 | 4 | 13.33 | 17 | 56.67 | 9 | 30.00 |
| p-tau217/Aβ42 | IJV | 3 | 10.00 | 27 | 90.00 | 3 | 10.00 | 8 | 26.67 | 19 | 63.33 |
| | MCV | 4 | 13.33 | 26 | 86.67 | 4 | 13.33 | 11 | 36.67 | 15 | 50.00 |
| p-tau181/Aβ42 | IJV | 10 | 33.33 | 20 | 66.67 | 3 | 10.00 | 14 | 46.67 | 13 | 43.33 |
| | MCV | 9 | 30.00 | 21 | 70.00 | 2 | 6.67 | 19 | 63.33 | 9 | 30.00 |
| Simoa | | Single-cutoff | | | | Two-cutoffs | | | | | |

| | | Negative | | Positive | | Negative | | Intermediate | | Positive | |
|---|---|---|---|---|---|---|---|---|---|---|---|
| | | No. | % | No. | % | No. | % | No. | % | No. | % |
| Aβ42/40 | IJV | 3 | 10.00 | 27 | 90.00 | 0 | 0.00 | 20 | 66.67 | 10 | 33.33 |
| | MCV | 11 | 36.67 | 19 | 63.33 | 2 | 6.67 | 16 | 53.33 | 12 | 40.00 |
| p-tau217 | IJV | 8 | 26.67 | 22 | 73.33 | 4 | 13.33 | 9 | 30.00 | 17 | 56.67 |
| | MCV | 8 | 26.67 | 22 | 73.33 | 4 | 13.33 | 6 | 20.00 | 20 | 66.67 |
| p-tau217/Aβ42 | IJV | 9 | 30.00 | 21 | 70.00 | 3 | 10.00 | 18 | 60.00 | 9 | 30.00 |
| | MCV | 6 | 20.00 | 24 | 80.00 | 4 | 13.33 | 15 | 50.00 | 11 | 36.67 |
